# Evaluating community knowledge, attitude, and practices toward implementation of telemedicine in Saudi Arabia

**DOI:** 10.1101/2023.09.27.23296248

**Authors:** Mustafa Mohammed Mustafa, Mohammed Al-Mohaithef

**Affiliations:** Department of Public Health, College of Health Sciences, Saudi Electronic University, Riyadh, Saudi Arabia

**Keywords:** Telemedicine, attitudes, cross-sectional, Riyadh city, COVID-19 pandemic, interactive medicine

## Abstract

**Background:** The urgent need to provide fast, low-cost, affordable, and high-quality medical services is one of the requirements of today’s modern world, which is characterized by a fast pace and search for the best services, especially regarding medical consultation and treatment. These demands have led to the invention of new ways that medical services can be provided to patients in their homes.

**Methods:** A cross-sectional study, conducted in February 2022 and ended in June 2023 with a representative sample of different categories of citizens. The study population included 281 (81.9%) males and 62 (18.1%) females (SD.385, mean 1.18). The study aimed to assess the knowledge, attitudes, and practices of Saudi citizens in the Riyadh region regarding telemedicine.

**Results:** The study revealed that only 30.3% of respondents were able to select the correct definition of the term telemedicine from three alternatives, which may reflect poor knowledge about telemedicine. A total of 19.5% of our participants said that they used telemedicine services only for medical consultations, while 13.7 said they used it for diagnosis and treatment. A total of 104 (88.9) out of 117 of the respondents believed that telemedicine has a positive impact on reducing costs and saving time.

**Conclusion:** The study concluded that the main barriers that limit the use of telemedicine services are a lack of confidence in accessing telemedicine, difficulty in communicating due to the use of different languages and dialects, and the fact that services are not suitable for uneducated people.

## Introduction

According to our internet search, all studies on telemedicine in the Kingdom of Saudi Arabia [KSA] were either review articles or only included physicians and, in some cases, university students [1, 2, 3]. This study included most of the community populations with their different specialties. Other studies on telemedicine were associated with COVID-19, and this factor may have some influence on the results given that people during the pandemic tried to avoid visiting health care services, gave attention and used other methods for medical consultation and treatment, such as telephone calls and online contact with their physicians [9, 11].

Saudi citizens are characterized by their widespread use of social devices, such as mobile phones and laptops, and are known to be knowledgeable about various applications and to have great confidence in these devices and the internet. Given the existence of an effective and fast internet service, the results of this study can be used to create positive attitudes, strengthen confidence about the reality of telemedicine, and convince citizens to adopt telemedicine, especially in light of the circumstances of the global COVID-19 pandemic, during which remote interaction has become paramount.

Developing the relationship of KSA citizens with telemedicine, which can lead to reduced pressure on health care facilities, a decreased equipment and operations budget, thus making the internet more useful by playing a role in promoting community health.

### Literature review

In the modern medical dictionary, telemedicine is simply defined as “the remote delivery of health care services” [4]. This means that health care services should be delivered to people in their homes when they adopt telemedicine. There are three common types of telemedicine, which include but are not limited to store-and-forward telemedicine and remote patient monitoring [5].

In Saudi Arabia, studies conducted in the past few years have revealed that the use of telemedicine has become widespread among health professionals in various hospitals [6]. An extensive review of the literature in the field of telemedicine in Saudi Arabia resulted in few publications on the identification of barriers and challenges inherent to telemedicine [7]. Another study was conducted in the KSA in 2021 to assess physicians’ knowledge and perceptions about, and willingness to use telemedicine in the Riyadh region of Saudi Arabia [8].

Physicians’ perceptions of telemedicine use during the COVID-19 pandemic in Riyadh, Saudi Arabia, were studied through a cross-sectional survey in 2022. The questionnaire was distributed among 500 physicians who employed telemedicine in their practice between October 2021 and December 2021. This study revealed that 70% (n=254) of responding physicians believed that telemedicine consultations are cost-effective. Regarding burnout, 4.1% (n=15), 7.5% (n=27), and 27.3% (n=99) of the doctors reported feeling burnout every day, a few times a week, and a few times per month, respectively [9].

The most interesting findings from a recent study reported four major concerns by various health professionals from four main hospitals in the KSA against the adaptation of telemedicine.

These concerns were mainly patient privacy, the high cost of equipment, the lack of suitable training and the lack of consultation between information technology experts and medical staff. All these stated factors are inconsistent with various published studies, and more studies should be conducted in this field to fill the gap between the theoretical and practical implementation of telemedicine in the field [9].

According to our web search, there are few studies conducted among everyday citizens except for two studies that were conducted among university students and faculty members, and such studies cannot satisfy the issue because university students and faculty members are recognized as being well educated and they do not represent the whole community. These studies revealed that both students and faculty members do not trust telemedicine and insist that face-to-face medication is essential, especially when they have severe symptoms or unfamiliar disorders.

Another study conducted among 314 responses aimed to study the factors affecting the adoption and acceptance of e-health in the context of Saudi Arabia. The study showed that usefulness and privacy significantly affected e-health adoption. Moreover, the perceived ease of use factor has an indirect effect on people’s perception of e-health services. Furthermore, the results show that system quality and trust are not influencing factors. [10].

Generally, telemedicine is the most preferred method for medical consultation and treatment because of the easy access to a network of doctors from the largest group of hospitals, and because the independence and autonomy of patients is increased by offering extra information and advice on medical issues.

## Material and methods

### Study Design and Setting

A cross-sectional study, conducted in February 2022 and ended in June 2023. The study designed to assess citizens’ knowledge, attitudes, and practices regarding telemedicine. Sociodemographic characteristics were measured using a well-constructed questionnaire that was adopted from the literature and modified to align with the research questions.

### Questionnaire Development

A well-constructed questionnaire that was adopted from the literature and modified to align with the research questions. The questionnaire gauged citizens’ knowledge, attitudes, and practices (6 items for each) and sociodemographic characteristics (6 items). Attitudes and experience questionnaires were rated using a 5-point Likert scale (strongly disagree to strongly agree). To ensure measurement consistency, the internal reliability of the questionnaire was measured using Cronbach’s alpha (α). Missing data were managed according to the type and magnitude of missing data while preserving data integrity.

### Study Sample

A sample of participants from organizations and governmental institutions was invited by email to participate in an online questionnaire. Other community groups were distributed into clusters and then investigated using the same questionnaire by personal contact, adopting opportunity sampling methods for each cluster. According to the latest statistics from the Saudi Statistics Authority in April 2021, the total number of Saudi citizens in Riyadh city was 27% less than 15 years ago (male: 1.633322, female: 1.500.195, total =3.138.517). The sample size consisted of 343 participants and was calculated using statistical equations at a 95.0% confidence level and a margin of error of 4.99%. The participants were selected randomly from all study populations. A representative sample size from all clusters was selected randomly. The total number of participants according to the statistical sample technique consisted of both males and females. All statistical tests adopted a significance level of 0.05.

### Ethics Statement

The study was conducted in accordance with the Declaration of Helsinki and approved by the Institutional Review Board and Ethics Committee of Saudi Electronic University. (SEUREC-220019-, 17.2.2022). Informed Consent Statement: “Informed consent was obtained from all subjects involved in the study.

### Data analysis

The responses recorded, validated and analyzed using appropriate statistical software (preferred SPSS 26). Descriptive analysis conducted to obtain variables distributions, central tendencies, frequencies, and proportions. Infernal analysis conducted to measure the magnitude of the Correlation between different variables and factors analyzed using appropriate statistics. Linear regression analyses utilized to assess the association between attitude and other variables. All statistical tests adopted a significance level of 0.05.

### Results

The characteristics of the study samples are shown in Table 1. A total of 343 participants agreed to complete the questionnaire online or accepted the paper questionnaire. The study population included 281 (81.9%) males and 62 (18.1%) females (SD.385, mean 1.18). The average age between 24 and 34 years was predominant for both males and females, with percentages of 33.8% and 35.5%, respectively. For level of education, the descriptive data showed that most males had bachelor’s degrees (128, 45.5%) and that most females also had bachelor’s degrees (25, 8.9%).

**Table 1.** The characteristics of the study population (sex, age, and education level).

| Sex |  | Male |  | Female |  |
| --- | --- | --- | --- | --- | --- |
|  |  | Count | % | Count | % |
| Age | 18–24 | 60 | 21.4 | 10 | 16.1 |
|  | 24–34 | 95 | 33.8 | 22 | 35.5 |
|  | 35–44 | 77 | 27.4 | 22 | 35.5 |
|  | 45–54 | 49 | 17.4 | 8 | 12.9 |
|  | Total | 281 | 100 | 62 | 100 |
| Education level | Post-graduate degree | 28 | 10.6 | 11 | 3.9 |
|  | Bachelor's degree | 128 | 45.5 | 25 | 8.9 |
|  | Secondary school | 86 | 30.6 | 18 | 6.4 |
|  | Intermediate school | 6 | 2.1 | 1 | 0.4 |
|  | Primary school | 23 | 8.2 | 5 | 1.8 |
|  | Other | 10 | 3.6 | 2 | 0.7 |

In Table 2. Regardless of sex, only 104 (30.3%) of the respondents selected the correct definition for the term (Telemedicine), and most of them had post-graduate education (31 (29.8%) PhD and master’s degrees holders or 50 (48.1%) bachelor’s degrees holders). Males represented more than 70% of respondents.

**Table 2.** Correct answer for the definition of telemedicine according to sex (descriptive).

|  |  |  | Education levels |  |  |  |  |  | Total |
| --- | --- | --- | --- | --- | --- | --- | --- | --- | --- |
|  |  |  | Post-graduate (PHD, Master's) | Bachelor's degree | secondary school | Primary school | Other | Intermedia te school |  |
| Selected the correct definition | sex | F | 8 | 9 | 4 | 0 | 0 | 4 | 25 |
|  |  | M | 23 | 41 | 7 | 1 | 3 | 4 | 79 |
|  | Total |  | 31(29.8%) | 50(48.1%) | 11(10.6%) | 1(1.1) | 3(2.9%) | 8(7.7%) | 104(30.3%) |

Detailed statistics in Table 3, showed that the mean difference between males and females was 2.347 in the degrees of freedom (342), and that the results were significant (.000) in the confidence interval (95%).

**Table 3.** Definition of telemedicine by sex.

| One-Sample Test |  |  |  |  |  |  |
| --- | --- | --- | --- | --- | --- | --- |
| Test Value = 0 |  |  |  |  |  |  |
|  | t | df | Sig. (2-tailed) | Mean Difference | 95% Confidence Interval of the Difference |  |
|  |  |  |  |  | Lower | Upper |
| How telemedicine is best defined | 38.160 | 342 | .000 | 2.347 | 2.23 | 2.47 |
| Sex | 56.744 | 342 | .000 | 1.181 | 1.14 | 1.22 |

The descriptive (frequency) analysis in Table 4 shows that only 118 (34.4%) of the respondents said that they had used telemedicine services before, while 225 (65.6%) had no practical experience with telemedicine.

**Table 4.** The use of telemedicine services.

|  |  | Frequency | Percent |
| --- | --- | --- | --- |
| Valid | Yes | 118 | 34.4 |
|  | No | 225 | 65.6 |
|  | Total | 343 | 100.0 |

Table 5 shows the effect of education on developing a positive attitude and practice regarding telemedicine. The differences were very significant when comparing the use of telemedicine services with the level of education (p value.000).

**Table 5.**
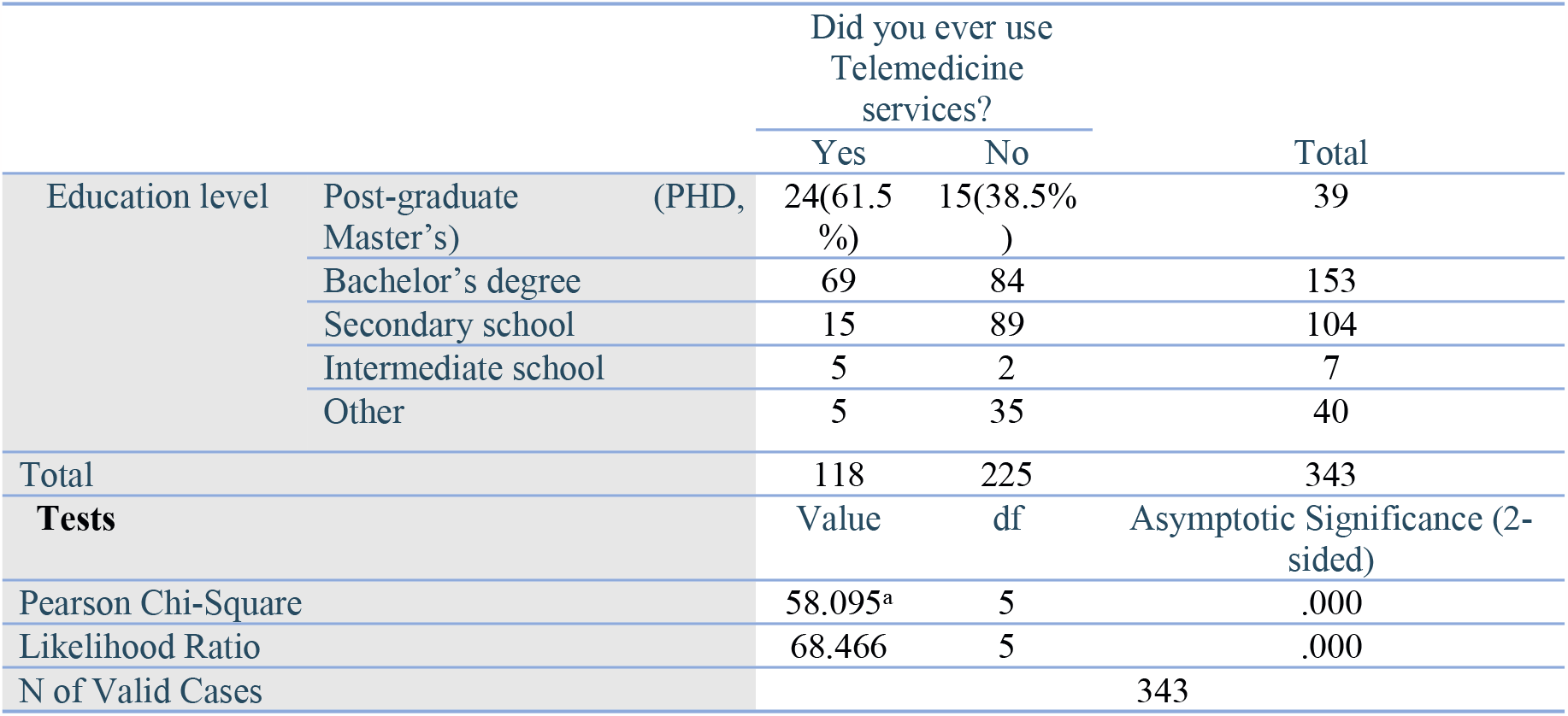
Use of telemedicine services cross tabulation with education level.

Table 6 shows that there were no significant differences when the use of telemedicine services was compared with sex, the level of education, and knowledge regarding the definition of the term telemedicine.

**Table 6.**
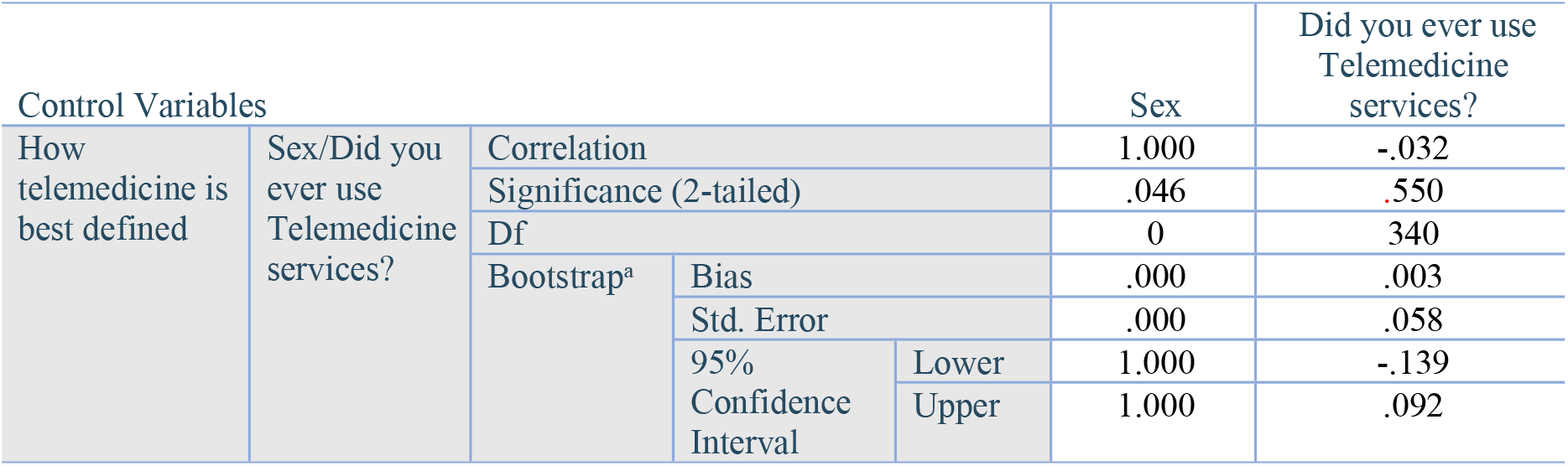
The use of telemedicine services cross tabulation with sex.

Table 7 provides information about the purposes of using telemedicine. A total of 19.5% of participants answered that they used the services for medical consultation only, while 13.7 said they used it for diagnosis and treatment.

**Table 7.** Purpose of using telemedicine services.

|  | Purpose | Frequency | Percent |
| --- | --- | --- | --- |
| Valid | Medical consultation only | 67 | 19.5 |
|  | Diagnosis and Treatment | 47 | 13.7 |
|  | Other | 4 | 1.2 |
|  | Total | 118 | 34.4 |

The study gave clear results about the opinions of the study group that used telemedicine services about its positive effects in terms of saving time and money. Most of the participants chose the levels very high, high, and average (Table 8).

**Table 8.** Positive impact of telemedicine on time and cost.

| Likert scale |  | Positive impact of telemedicine on time |  | Positive impact of telemedicine on cost |  |
| --- | --- | --- | --- | --- | --- |
|  |  | Frequency | Percent | Frequency | Percent |
| Valid | Very high | 22 | 6.4 | 21 | 6.1 |
|  | High | 45 | 13.1 | 37 | 10.8 |
|  | Average | 47 | 13.7 | 48 | 14.0 |
|  | Low | 2 | 0.6 | 6 | 1.7 |
|  | Very low | 1 | 0.3 | 6 | 1.7 |
|  | Total | 117 | 34.1 | 118 | 34.4 |
| Missing data |  | 1 | 0.84 | 0 | 0 |

Approximately 37% of the respondents gave a high score to the negative effect of telemedicine on relationships between patients and doctors, compared with 15% who chose a low effect (Table 9).

**Table 9.** The effects of telemedicine on the doctor-patient relationship.

| Likert scale |  | Frequency | Percent |
| --- | --- | --- | --- |
| Valid | Very high | 42 | 12.2 |
|  | High | 127 | 37.0 |
|  | Average | 107 | 31.2 |
|  | Low | 52 | 15.2 |
|  | Very low | 15 | 4.4 |
|  | Total | 343 | 100.0 |

The main barriers that limit the use of telemedicine services are clarified by the respondents in Table 10, as they chose a level (high) on a Likert scale for the first three options (Lack of confidence in accessing telemedicine, Difficulty in communicating due to different languages and dialects, The service is not suitable for uneducated people), while many of them believe that communication and internet services will not be a major obstacle to them when they think of using telemedicine.

**Table 10.**
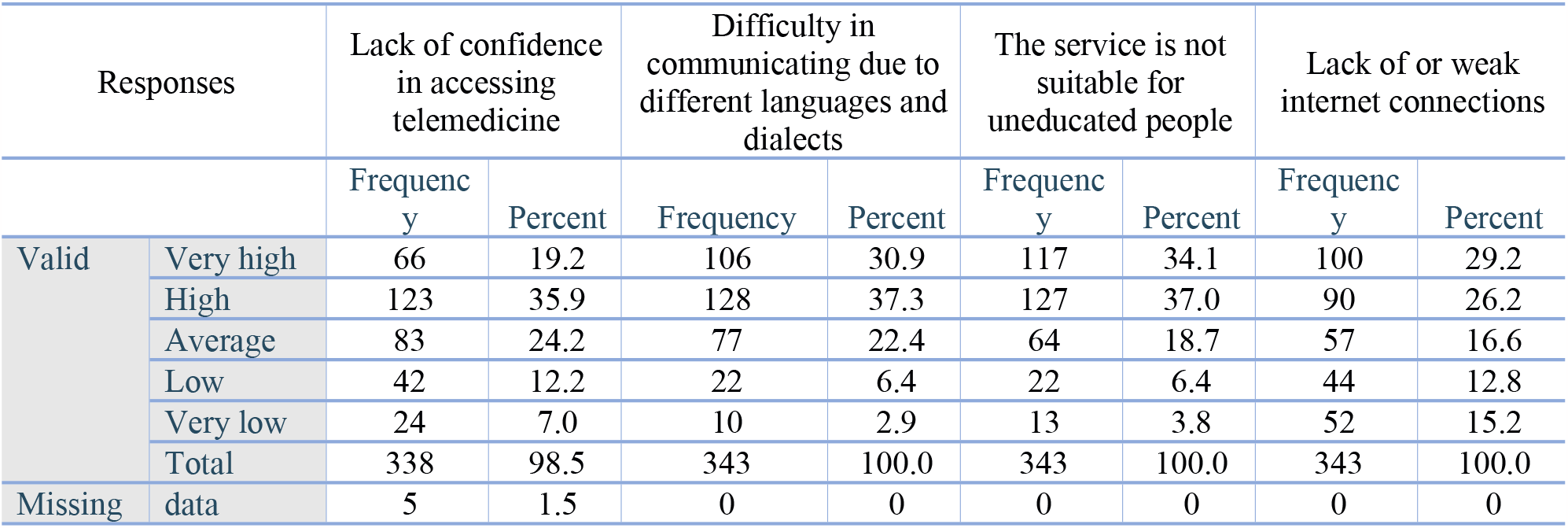
The main barriers to using telemedicine services.

| Responses |  | Lack of confidence in accessing telemedicine |  | Difficulty in communicating due to different languages and dialects |  | The service is not suitable for uneducated people |  | Lack of or weak internet connections |  |
| --- | --- | --- | --- | --- | --- | --- | --- | --- | --- |
|  |  | Frequency | Percent | Frequency | Percent | Frequency | Percent | Frequency | Percent |
| Valid | Very high | 66 | 19.2 | 106 | 30.9 | 117 | 34.1 | 100 | 29.2 |
|  | High | 123 | 35.9 | 128 | 37.3 | 127 | 37.0 | 90 | 26.2 |
|  | Average | 83 | 24.2 | 77 | 22.4 | 64 | 18.7 | 57 | 16.6 |
|  | Low | 42 | 12.2 | 22 | 6.4 | 22 | 6.4 | 44 | 12.8 |
|  | Very low | 24 | 7.0 | 10 | 2.9 | 13 | 3.8 | 52 | 15.2 |
|  | Total | 338 | 98.5 | 343 | 100.0 | 343 | 100.0 | 343 | 100.0 |
| Missing | data | 5 | 1.5 | 0 | 0 | 0 | 0 | 0 | 0 |

## Discussion

Although our study was conducted during the COVID-19 pandemic, it seems that it had no significant effects on the study results, perhaps because the questions were open. There was also no indication that the responses were related to a specific temporary health condition (COVID-19), but there may still be some undetected effects.

The study aimed to assess the knowledge, attitudes, and practices of Saudi citizens in the Riyadh region regarding telemedicine. A total of 343 participants agreed to complete the questionnaire online or accepted the paper questionnaire. The study population included 281 (81.9%) males and 62 (18.1%) females (SD.385, mean 1.18). The average age between 24 and 34 years was predominant for both males and females, with percentages of 33.8% and 35.5%, respectively. For level of education, the descriptive data showed that most males had bachelor’s degrees (128, 45.5%) and most females also had bachelor’s degrees (25, 8.9%).

Our study revealed that only 30.3% of respondents were able to select the correct definition of the term telemedicine from three alternatives, which may reflect poor knowledge regarding telemedicine. The results contrast with the results of a study conducted in Saudi Arabia in 2019 among the Saudi population; most of the participants (70.0%) were familiar with the term

“telemedicine”. The only difference between the aforementioned study and ours is that most of our study population was male (81.9%), while females were prevalent (73.9%) in the above study [11]. On the other hand, the results of our study are similar to the results of the statistics, which indicated that 21%, more than half of the participants, were unaware of the services that telemedicine can provide [12]. With regard to the sex, statistics in Table 3, showed that the mean difference between males and females was 2.347 in the degrees of freedom (342), and that the results were significant (.000) in the confidence interval (95%).

The descriptive analysis of our study shows that only 118 (34.4%) of the respondents used telemedicine services, while 225 (65.6) had no practical experience with telemedicine. This result is similar to the results of a previous study conducted in the KSA in 2021, which showed that only 33% of the participants had used telemedicine services systems before [11].

Education level has clear effects in developing a positive attitude and practice towards telemedicine, and the differences were significant when comparing the use of telemedicine services with the levels of education (24 (61.5%) and 15 (38.5%) for post-graduate degree holders).

The study revealed that there were no significant differences when the use of telemedicine services was compared with sex and knowledge regarding the definition of the term telemedicine.

A total of 19.5% of our participants said that they used the services for medical consultation only, while 13.7% said they used it for diagnosis and treatment. This result is different from results from a study conducted in December 2020 in Saudi Arabia, which showed that 305 (77.8%) were conducting routine (live) patient consultations since the start of the pandemic (the period being approximately two months); 228 (58.1%) had used some form of telemedicine (other than standard phone calls) during the COVID-19 pandemic [13].

A total of 104 (88.9) out of 117 of the respondents believed that telemedicine has a positive impact on reducing costs and saving time. Similarly, some preceding studies have suggested that telemedicine is an accessible and convenient tool that decreases wasted time on travelling and costs [14, 15, 16, and 17]. The results are comparable with a study indicating that telemedicine can reduce unnecessary outpatient visits (87.5%) [13].

Approximately 37% of the respondents gave a high score to the negative effect of telemedicine on relationships between patients and doctors, compared with 15% who chose a low effect.

Our study concluded that the main barriers that limit the use of telemedicine services are a lack of confidence in accessing telemedicine (19.2%), with 35.9% of respondents giving it a very high score and 24.2% giving it an average score. Difficulty in communicating due to different languages and dialects was also one of the main barriers, with scores of 30.9%, 37.3%, and 22.4% for very high, high, and average, respectively. The service is not suitable for uneducated people, while many of them believe that communication tools and internet services will not be a major obstacle to them when they think of using telemedicine. In a similar study, the results showed that technological limitations (66.6%) and concerns about diagnostic reliability (66.1%) were the main barriers [13]. Other barriers, such as provider-patient relationships, medical liability, prescription of controlled substances and others, were mentioned in published scientific papers in 2020 [18]. In the KSA study, based on 905 respondents conducted in 2016, 11 barriers were highlighted to be most significant according to the KSA context. The findings of this study show that the top three influential barriers to the adoption and implementation of telemedicine by health care facilities decision-makers are the availability of adequate sustainable financial support, ensuring conformity of telemedicine services with the core mission, vision, needs and constraints of the health care facilities, and the reimbursement for telemedicine services [19].

## Conclusion

Telemedicine has a good opportunity to provide fast, low-cost, affordable and high-quality medical services in Saudi Arabia, either during certain health conditions (COVID-19) or other epidemics or even under normal situations. More efforts should be made to provide health care institutions and practitioners with technical equipment and training. Furthermore, improving knowledge regarding telemedicine among the population can develop positive perceptions and good attitudes. Other barriers must be taken into account.

## Data Availability

All relevant data are within the manuscript and its Supporting Information files.

## Acknowledgments

The authors are grateful to the Deanship of Scientific Research at Saudi Electronic University, KSA. Thanks, and appreciation should be extended to all participants who took the time to fill out the questionnaire for data collection.

